# Competing risks multi-state model for time-to-event data analysis of HIV/AIDS: A retrospective cohort national datasets, Ethiopia

**DOI:** 10.1101/2023.10.25.23297525

**Authors:** Tsegaye Hailu Kumsa, Andargachew Mulu, Adane Mihret, Zeytu G. Asfaw

## Abstract

**Introduction:** When HIV/AIDS patients die from opportunistic infections that cause death to happen more quickly, competing dangers are present. The objective of the current study was to estimate the probability of HIV/AIDS patients dying from competing illnesses such as tuberculosis, diarrhea, other opportunistic infections, and unidentified infections.

**Methods:** All regional states, including the administrative cities of Addis Abeba and Dire Dawa, a retrospective cohort research was carried out between November 2019 to March 2020. There were 39590 HIV-positive individuals considered. We used competing risk models with a time-to-death horizon of 1212 for the total number of HIV-positive people. A competing event was thought to be death from various causes.

**Results:** Out of the total 1212 deaths, 542(44.7%) died competing with other opportunistic infection (TE-Esophageal Candidiasis, TO-oral, CT-CNS Toxoplasmosis, CM-Crypotococcal Meningitis…), 421 (34.7%) died due to tuberculosis and the remaining death were unknown/Not specified infection 222(18.3%) and diarrhea 27(2.2%). Rates of mortality caused by tuberculosis, competing with other opportunistic infection, diarrhea and unknown/Not specified were 3.5, 4.5, 0.2 and 1.8 per 1000 person-months, respectively. Responding to combined Antiretroviral Treatment (cART) 6 months after initiation, receiving Pneumocystis Pneumonia (PCP) prophylaxis, and higher CD4 count at diagnosis reduced the hazard of tuberculosis, other opportunistic infection and unknown and diarrhea causes of death. However, older age, late HIV.AIDS diagnosis, and the last HIV/AIDS WHO clinical stages increased the hazard of tuberculosis and other opportunistic disease mortality. Additionally, male gender, older age and last HIV clinical stages increased the mortality HIV/AIDS patients.

**Conclusion:** The findings of this study demonstrated that TB, an opportunistic infection, was the primary cause of death in HIV/AIDS patients, despite the presence of several competing risks, such as diarrhea, other infections, and an undetermined or unclear cause. It’s important to use effective techniques to quickly detect those who have HIV or AIDS and provide them with care and treatment to increase their chances of surviving.

## Introduction

According to estimates, HIV has affected 40.1 million people worldwide and is still transmitting in all nations, with 25.6 million of those cases occurring in the African region [1, 2]. HIV infection affected 1.5 million individuals in 2021, and 650000 people passed away from HIV-related causes (1, 3). In 2022, there will be about 10421 fatalities each year in Ethiopia across all age groups (4).

Antiretroviral therapy (ART) has increased the long-term survival rate of HIV-infected patients and transformed their outlook from a terminal sickness to a protracted and treatable condition. But this depends on a number of risk variables, such as if the HIV patient has TB, diarrhea, other opportunistic illnesses, or an unknown infection. As a result, rather than the virus itself, an HIV/AIDS patient died from TB and opportunistic illness that were growing more common in the population. In order to develop effective intervention strategies, improve patient care, and minimize HIV patient mortality as much as possible, causes of death (CoD) among HIV-infected patients must be evaluated (5). Human immunodeficiency virus (HIV) patients receiving antiretroviral therapy (ART) were the subjects of cohort studies, where participants might go through a variety of circumstances. One must consider the possibility of opportunistic infectious disease while examining the time to death process in HIV positive individuals, for example. Understanding the prognostic variables affecting HIV patients’ long-term survival is necessary (6, 7). In addition to the risk of dying from tuberculosis, HIV/AIDS patients also run the risk of dying from other infections or diarrhea. Competing risks arise when participants can witness one or more events, they ‘compete’ with one another, and the occurrence of one event could make it impossible to witness the other events or change the likelihood of their occurrence. It is customary to achieve this by carrying out separate studies for each end point and the intermediate events, but this is insufficient because it disregards the connections between these events. Multistate models (MSM) are a natural technique to model such complex systems in this context (8).

A particularly helpful tool for answering a variety of survival analysis concerns that traditional models are unable to address is the MSM framework (9). Figure 1 displays the two simplest but most typical instances. Although the current HIV/AIDS study is primarily focused on deaths caused by tuberculosis, some patients may experience many competing causes of death. Although fatalities from other causes occur in place of tuberculosis deaths, the event of interest is a death from tuberculosis. Comparative risk model assessed cause-specific mortality in patients on highly active antiretroviral treatment (HAART) from tuberculosis, diarrhea, other opportunistic infections, and unknown/unspecified causes (10–12). In patients who died from opportunistic infections while on HAART, there are many international literatures that quantify death from other causes (13–17).

However, relatively few researches were carried out in Ethiopia, for example, at the Gonder, Dilla, and Pawai hospitals. Their attention was on HIV patients who were lost to follow-up who presented competing risks. In this study, death-competing risks from tuberculosis, diarrhea, other opportunistic infections, and unidentified infections are examined using national statistics. Such research could be a useful contribution to the Ministry of Health’s efforts to develop appropriate intervention strategies and enhance patient care. Reduced mortality from AIDS-related opportunistic infections is also very helpful for HIV-positive patients.

The Kaplan-Meier estimator and Cox proportional hazard model are the traditional classical survival analysis techniques for time-to-event data (18, 19). These techniques work well in investigations where there is just one primary event type of interest. The aforementioned techniques might not fully reveal the relationship when there are several occurrences of interest. Methods that can reveal the underlying connection between the variables, the intermediate outcomes, and the outcomes of interest are required in these circumstances. To expand on tried-and-true techniques, multistate models offer a versatile and expansive framework (20). The competing risks model, which is a special case of a multi-state model and is a cause specific hazard, sub-distribution hazard, and flexible parametric hazard model, was used in this study. Because of this, the main goal of this article was to estimate the risk of death for HIV patients using ART from tuberculosis, diarrhea, other opportunistic infections, and unidentified infections in the lack of covariates and the existence of competing hazards. Finally, the cumulative Incidence Function (CIF) was utilized to predict the likelihood of mortality and the cause specific hazard, sub-distribution hazard, and flexible parametric hazard model were employed to evaluate the covariate impacts on CIF.

## Material and Methods

**Source population:** The source population of this study (record review) constituted all medical records of HIV/AIDS patients who initiated and were enrolled on anti-retroviral therapy at all health facilities in Ethiopia.

**Study population:** The study population is classified from health facility and individual perspectives as follow:

### Inclusion criteria

Health Facilities: Governmental, non-governmental, private or uniformed hospitals and health centers that have been providing ART service for at least one year prior to the study period. Records for review: records of individuals diagnosed with HIV/AIDS and at least one year since they have initiated ART at time of data collection.

### Exclusion criteria

Health centers having <100 patients whoever started ART were excluded so as to get adequate records and be cost effective while moving to that site. Catchment Health center which are more than 30 km from the mentoring hospital or Center (Zonal town).

## Sample size determination and sampling procedure

### Sampling techniques

Standard probability sampling technique was applied to select nationally representative sample from the study population, and region was the domain of analysis. The primary sampling units are ART patient charts in the hospitals and catchment health centers providing ART.

### Sampling

Multi-stage clustered sampling method was used to select study facilities and participants. The steps followed to select the regions, facilities, and allocate ART sample to each facility is depicted in to three strata:

### Sample size determination

This study was more or less a replica of the second round ART program effectiveness assessment done back in 2011. Hence, the sample size was derived purposively by using the cohort sampled population size from the previous study of around 40,000 ART patient charts to be reviewed for comparison sake. Finally, at the end of the data collection we ended up retrieving and reviewing a total of 39,590 medical records, which includes all 5,802 pediatric ART patient records in the facilities.

## Data Collection Tools and Procedures

The instrument measures age at baseline, gender, baseline WHO stage, baseline functional status, and outcome (alive and on treatment, transferred out, lost to follow-up, known dead). Moreover, achieved ART outcomes explained by quality of life such as CD4 count, weight, functional status, and 7 years survival rates by strata and by different time points were also assessed. Likewise, it also measured self-reported adherence to treatment and care, dropout rate, drug pick up behavior, and retention on 1st line regimen. It detected problems in the flow of the questions, gauge the length of time required for interviews, and identify problems in the understanding of terms and concepts.

### Pretest

The instrument was pretested and piloted during the training of the data collectors, and the modification and amendment were done by all the trainees’ feedback and valuable inputs. It At National level, the NAPES team was basically structured into nine routes established based on the main transportation roads in the country. In late-January, three additionally new sub routes were added to assist the existing nine routes. Each route consisted on average of 5 - 7 hospitals, and the period of stay at each facility was ranging from 5 −10 days based on the sampled ART patients/case load in that facility. The data collection comprised of gathering GPS coordinates, facility level information by interviewing the facility head, and data extraction of ART patient chart.

## Data Quality Assurance

Quality assurance began with the recruitment of data collectors and team leaders with a health background. Data collectors took pre- and post-test to assess their learning and knowledge of the assessment guidelines and standards for data collection. An intensive 8 days training was given to all field team. Each data collector was given a hard copy manual of the assessment guidelines for reference in the field. The central team, performed data quality checks, followed by a data quality control Skype call between the whole team to discuss any data collection or quality issues. Also, based on teams’ feedback, frequent modification of the Redcap tool was done to make the tool user friendly and ensure quality.

### Data preparation

#### Predictors variables

In this survey, a retrospective cohort study design was employed. It was a national study involving nine regional states and two city administrations from November 2019 to March 2020. The study participants were selected randomly from patient registers and medical data those started ART. Age, weight, hemoglobin, sex, marital status, educational level, region, and type of health facility were among the demographic data obtained. The retrospective record review also revealed trends of ART treatment regimen change, adherence and TB status were also determined from the retrospective record review also revealed. The patient has had tests for opportunistic infections, TB status, liver enzyme, Hepatitis B and C, and family planning counseling. Additionally, each visit includes a CD4 count and viral load measurements.

#### Outcome variable

The primary endpoint for this study was mortality from AIDS related tuberculosis, diarrhea, other opportunistic infection and unknown/not specified causes. The study data collector recorded the causes of death using tablet and in cases where inadequate information is provided to determine the causes of death, the study coordinator contacts personnel at the study site for further information. Patients are classified as having an unknown or not specified cause of death if no further information is obtained.

### Multi-state models

A multi-state model is a model for time-to-event data in which all individuals start in one or possibly more starting states (e.g. HIV patient) and eventually may end up in one (or more) absorbing or final state(s) (e.g. TB or Diarrhea or other Opportunistic infection disease or unknown[1]. Some individuals are censored before they reach an absorbing state. Competing risks models are a sub-category of multi-state models: they have one starting state, at least two absorbing states and no intermediate states (see Fig. I) left panel [2]. Their transitions are often indicated as causes of failure. An example of a multi-state model is illustrated in Fig I right panel shown.

**Figure I.**
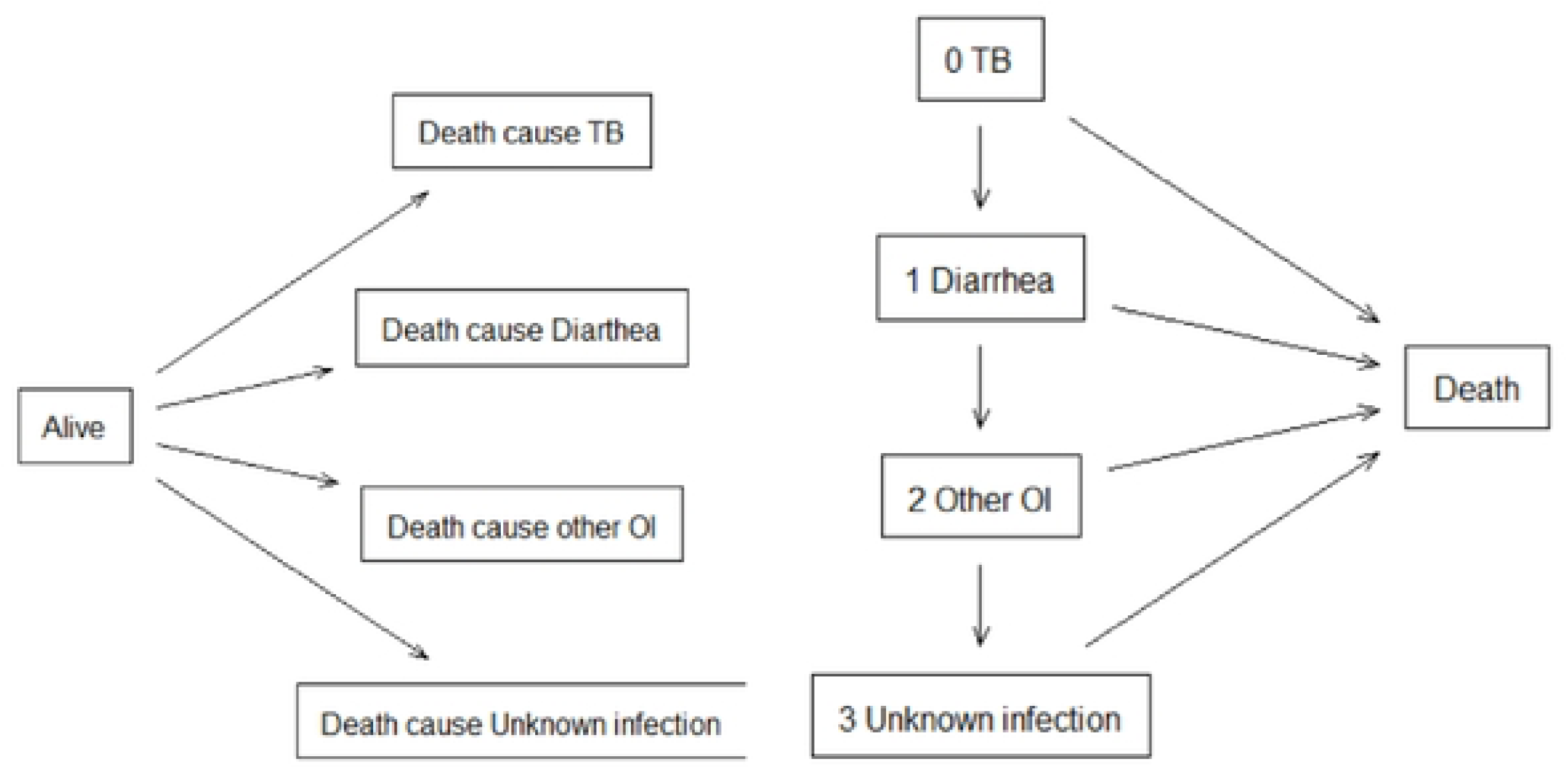
Title competing risks model with and death as causes of failure (left panel). Fig I legend Illness-death model with an initial state of illness state and a death state (right panel)

### The competing risks multistate model

A special kind of multistate model called competing risk analysis seeks to accurately predict the marginal probability of an event in the presence of competing events[3] When analyzing the marginal probability for cause-specific occurrences, the Kaplan Meier product-limit technique has a tendency to yield estimates that are erroneous since it is not designed to take into account the competing nature of numerous causes to the same event[4]. Cumulative Incidence Function (CIF) has been used to predict the chance of death due to competing risk. In this competing risk analysis, the cause specific hazard model and the sub distribution hazard model were utilized to analyses the covariate impacts on cause specific hazard[5]. A flexible parametric hazard model was also used to determine the importance of covariates[6] and the outcomes of these three strategies are then compared.

### 3.1 Hazard-based regression models

#### Cause-specific hazard regression

The most commonly used regression model for analyzing event-time data is the Cox proportional hazards model[7]. In the presence of competing risks, the standard Cox proportional hazards model is not adequate because the cause-specific Cox model treats competing risks of the event of interest as censored observations [8] In addition, the cause-specific hazard function does not have a direct interpretation in terms of survival probability[9]. An adaptation of Cox regression requiring data augmentation has been proposed. With k competing events, the data for each patient are duplicated k times, one row for each type of failure; then, k-1 indicator variables are created for identifying whether a certain event has occurred. A stratified Cox regression could also be applied to allow non-proportional hazards[10].

To make complete inference, we performed cause-specific hazard model and cause-specific hazard functions [2, 11–13] are defined as:

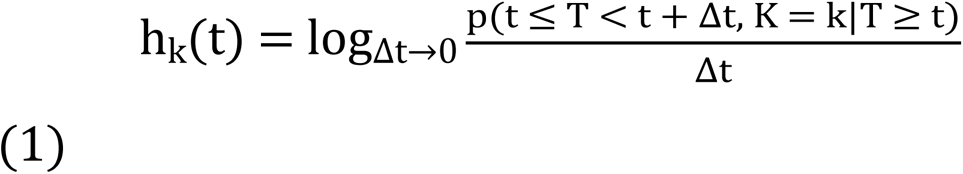

Where T be the survival time and K be the cause of death (k=1, 2, 3, 4).

The cumulative cause-specific hazard is written as:

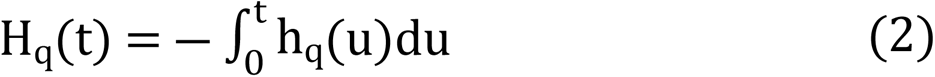

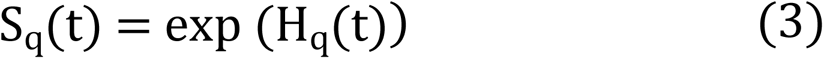

Here we assume that the event of interest is death of AIDS patient due to the opportunistic infection TB (indexed 1, 2, 3, 4), coded Tuberculosis (as the event of interest), diarrhea, other opportunistic infections and unknown causes respectively. Fig II tell us the model competing risk HIV/AIDS patients on ART.

**Figure II.**
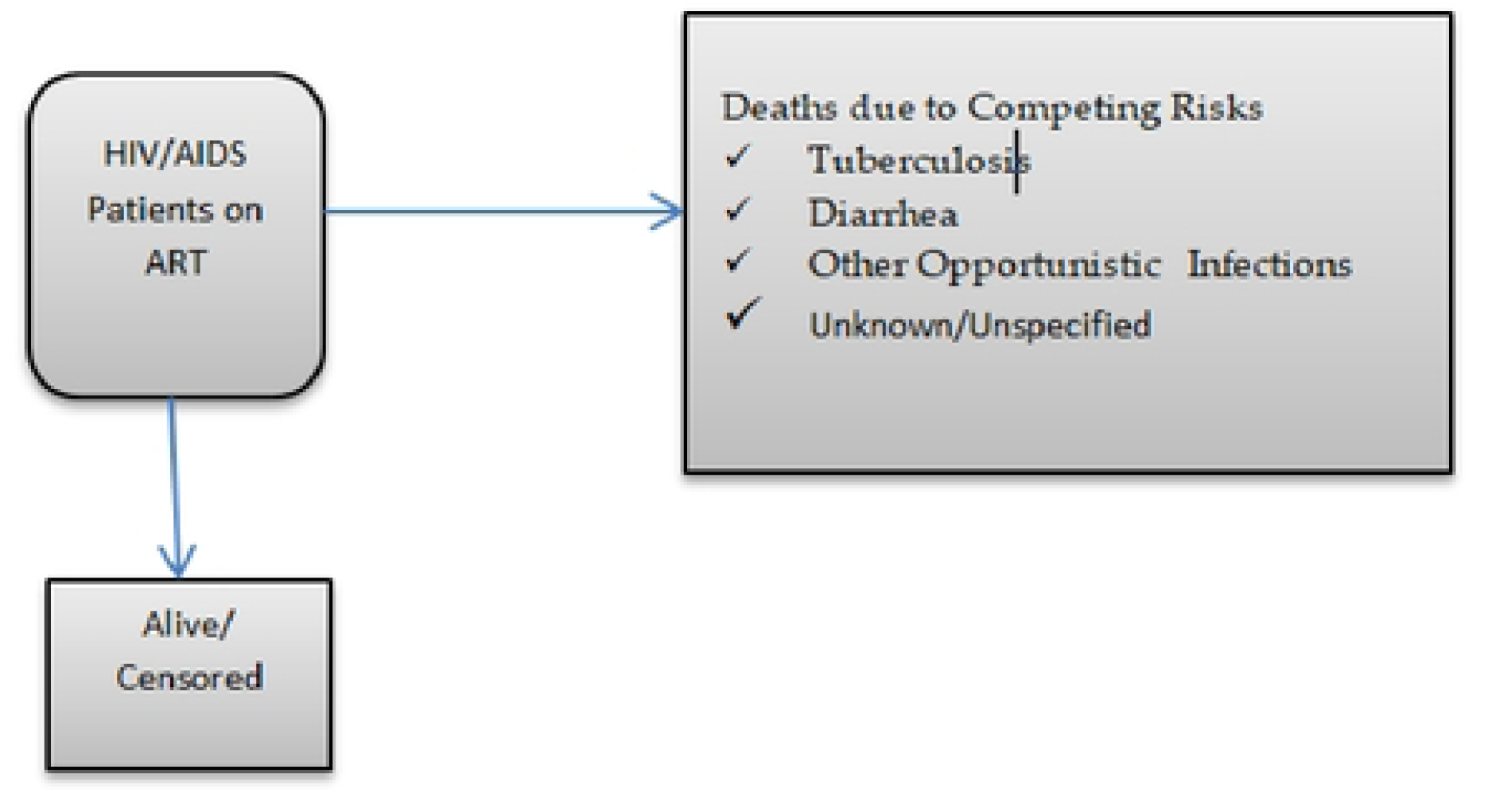
Title Competing Risks Model for HIV/AIDS Patients on ART.

#### Sub-distribution hazard regression—the Fine and Gray model

The Fine-Gray[9] sub-distribution hazard model is frequently used in the tuberculosis related diarrhea, Other opportunistic infection and unknown infections cause death to estimate subject-specific probabilities of the occurrence of an event of interest over time in the presence of competing risks. The sub distribution hazard is a key concept in this approach, and it is defined as the hazard of failing from a given cause in the presence of competing events, given that a subject has survived or has already failed due to different causes. We can write the sub-distribution hazard[5] for cause as

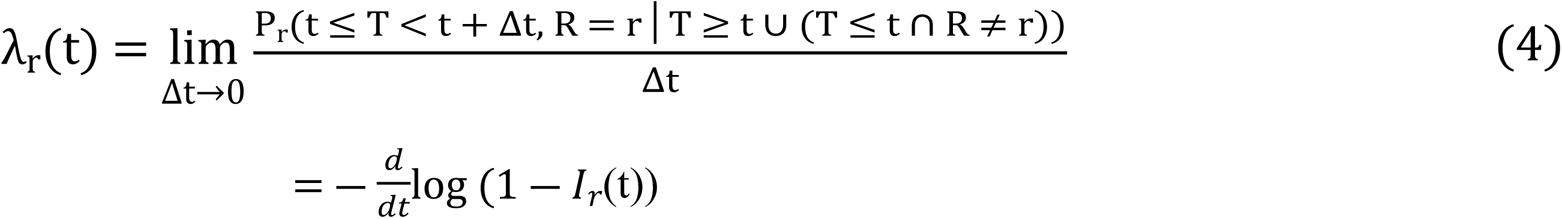

Where *I_r_*(t)) = Pr(T ≤ t, R=r) is the CIF for cause r (r =1, 2, 3).

A semi parametric proportional hazards model for the sub-distribution hazard of cause r for a subject with covariate vector X as follows

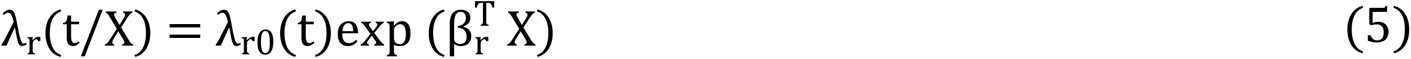

where λ_r0_(t) is the baseline subdistribution hazard of cause r, and β_r_is the vector of coefficients for the covariates. Estimation for this model follows the partial likelihood approach used in a standard Cox model.

#### Flexible Parametric Proportional Hazard Model

The flexible parametric model was first developed for use with censored survival data (6). The log cumulative hazard variant of a Weibull model was picked since it is the most prominent parametric survival model with both proportional hazards and accelerated failure time model interpretations. A Weibull distribution’s survival function is

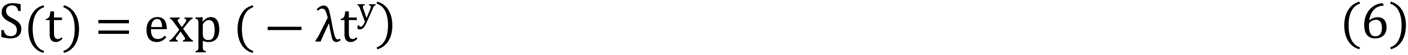

Transforming this to the log cumulative hazard scale, we get

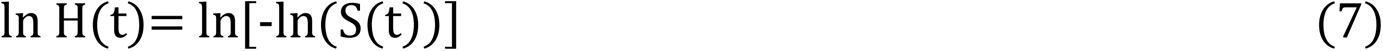

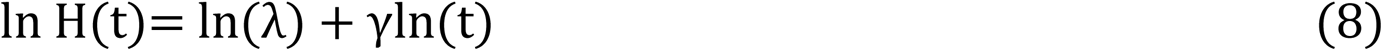

This is a linear function of log time, now adding covariates X in this model, we have

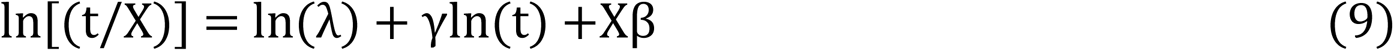

Where β are the covariate coefficients, but instead of assuming linearity with ln(t), adopt a flexible parametric method that employs restricted cubic splines for ln(t)(14). Under the premise of proportional hazards, covariate effects can be seen as log hazard ratios. Transforming the model parameters yields the survival and hazard functions.

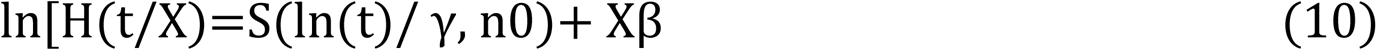

One of the primary advantages of the flexible parametric technique is the ease with which time dependent effects can be fitted.

## Result

In the present study, we have considered 39590 HIV patients. Out of the total HIV patients, 1212 were died during the period of observation from November 2019 to March2020. The characteristics of these people as at the time of their death are shown in Table 1. Out of the total, 34.7% were died due to TB; 44.7% were died of other opportunistic infection; and the remaining 18.3% and 2.2 % were died of unknown/Not specified and diarrhea respectively (Table 1 and Fig 1). At the time of death, the mean age of the HIV patients was 35±14.6 with mean weight of 48.7kg and 11.9±6.3 Hemoglobin. The majority of deaths nearly 80% were between WHO stage 2 and stage 4 and 639(67.33%) were CD4 count cell and the remaining 19.9% were stage1.The majority (73.3%) was divorce or widowed and the remaining 18.3% were single. Similarly, the mean weight was 48.7Kg and hemoglobin mean was 11.9±6.31.

**Table 1:** Title Characteristics of the respondents at death (N= 1212)

**Fig 1:**
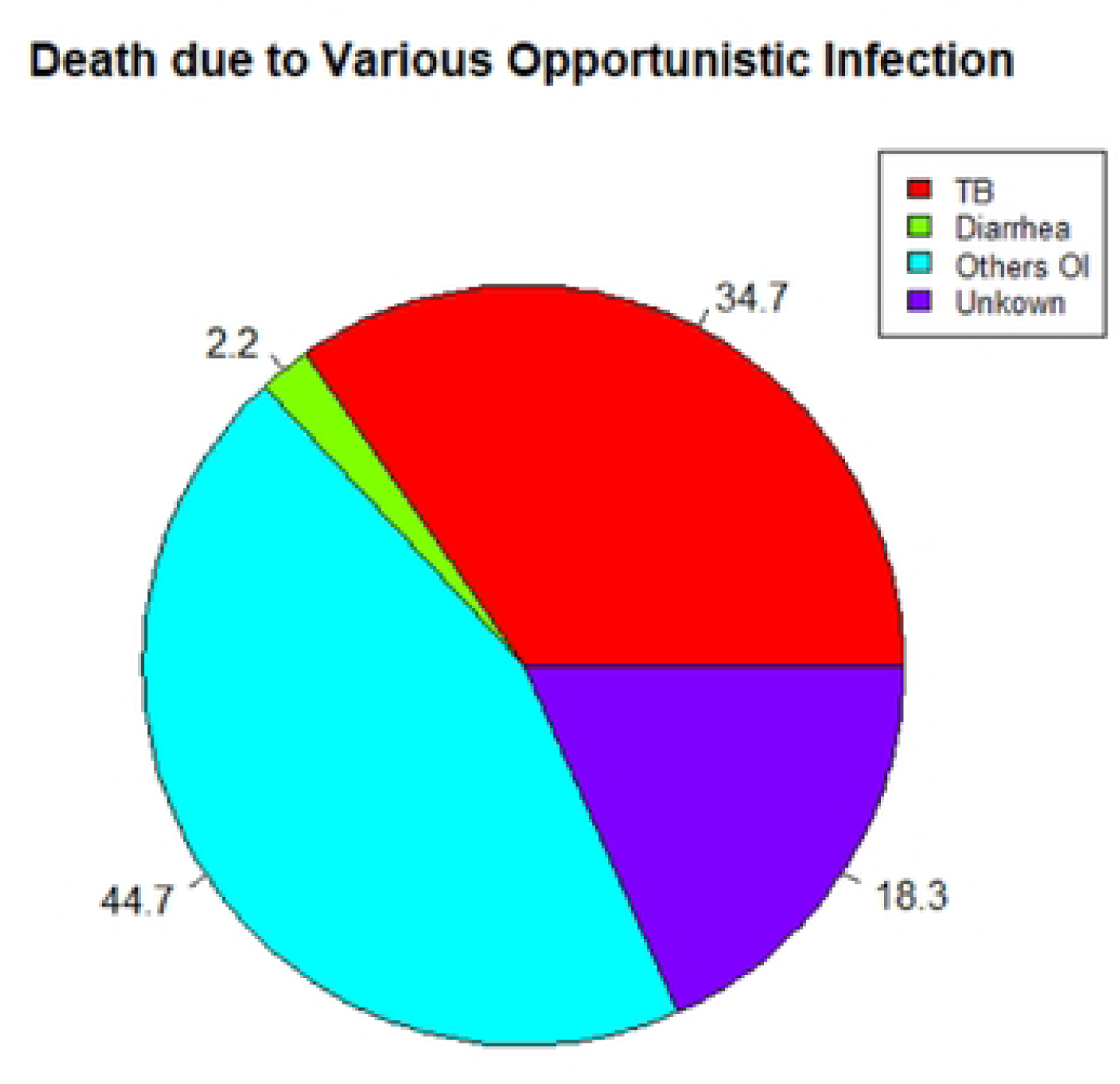
Title Death due to various opportunistic infection.

## Survival analysis

Male patients had better survival than female HIV patients for the first 45 months and after 75 months than female but female were better than male 46-75 months during the follow-up periods (Fig 2A). In figure 2B, patients had nearly high risk of probability of death due to other Opportunistic Infection and Diarrhea in the first 30 months but probability of death due to unknown were lower as compared to the others causes. After 40 months, Diarrhea had lower risk as compared to the others opportunistic infection and unknown infections.

**Fig 2.**
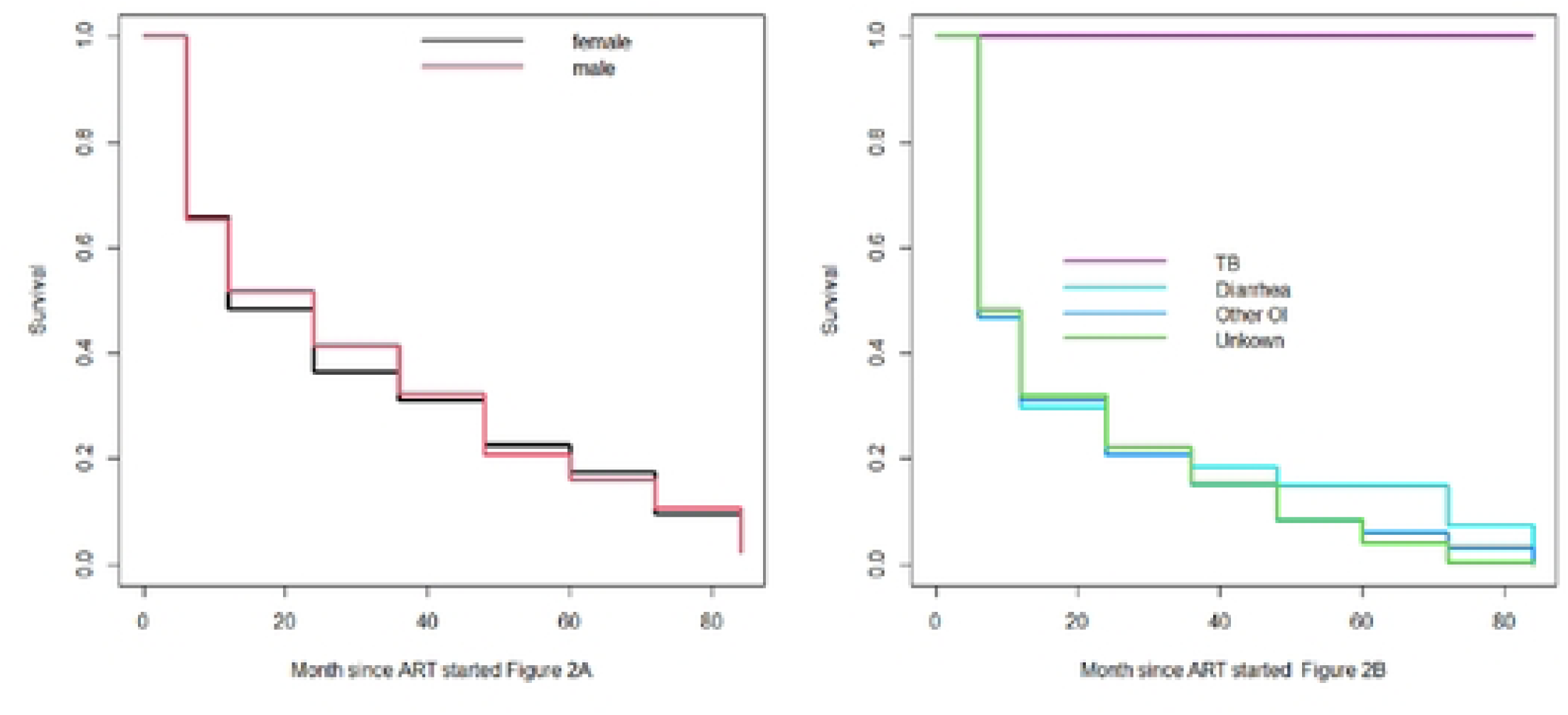
Title Kaplan-Meier curve. Fig 2 legend Fig.2A:Kaplan-Meier curve by male and female and Fig.2B: Kaplan-Meier curve by Tuberculosis, Diarrhea other opportunistic disease and unknown.

## Non-parametric comparison of cumulative incidence function (CIFs)

HIV/AIDS patients have higher risk of death from other opportunistic infection and followed by death from TB, Unknown and Diarrhea causes. As the time goes from 6-50 months, Male patients have slightly higher risk of death from Other Opportunistic Infection but from 50-84month Female patients were higher risk of death as compared to male. But in the causes (OI and TB) male had higher risk of death than female. Similarly, there is no significant difference in mortality risk from all causes of death for male versus female patients, since p-values (p= 0.48, 0.18, 0.21, and 0.75: TB, Diarrhea, other Opportunistic Infection and Unkown respectively) (Fig 3A).

**Fig 3.**
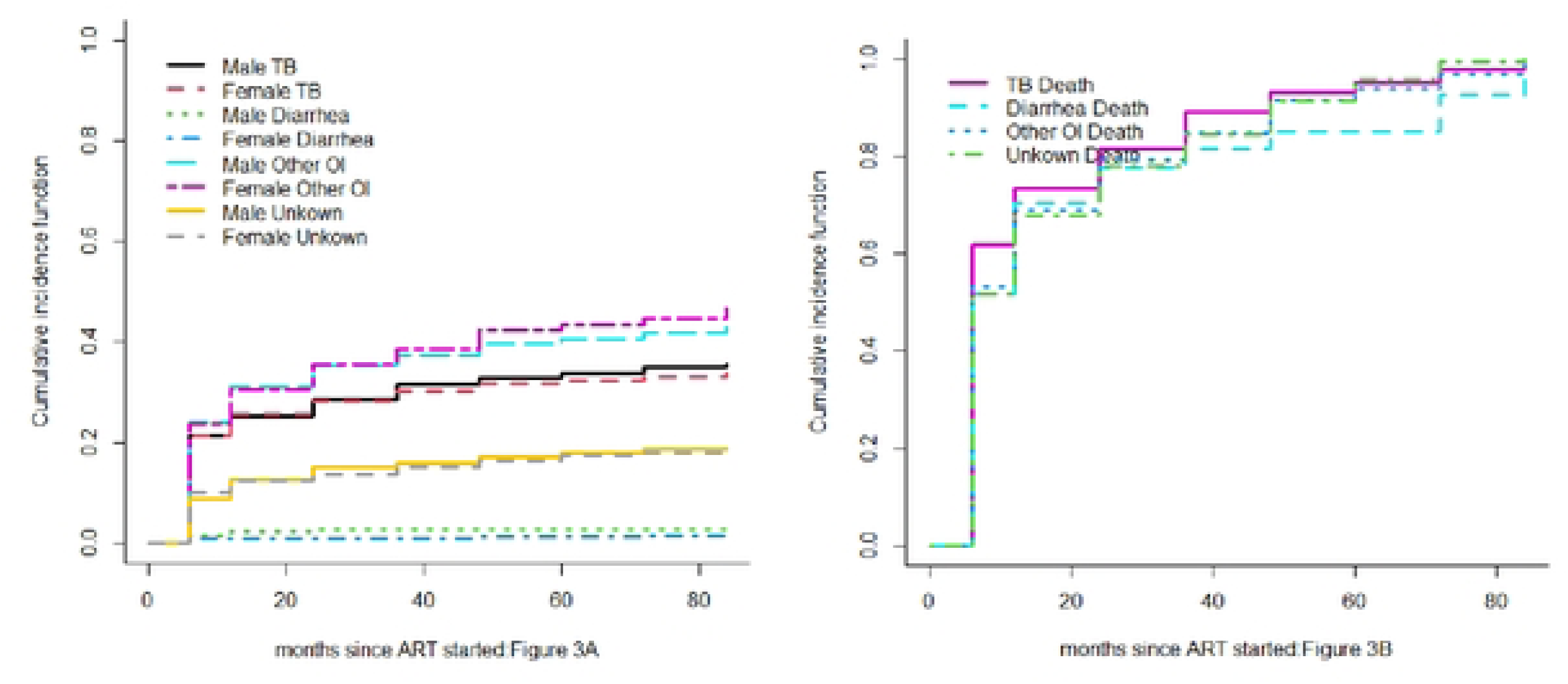
Title Non-parametric estimates of cumulative incidence. functions Fig 3 legend Fig3Adeaths from TB (status==1) Diarrhea (status=2), other Opportunistic Infection (status=3) and unknown causes (status=4).Each outcome is compared between male and female patients. **Fig 3B**: Estimate cumulative incidence in the presence of TB death, Diarrhea, Other OI & Unknown

Fig 3B above showed that the death from tuberculosis, Diarrhea, other opportunistic infection and unknown causes. From 6 month to 60 months, TB had higher risk of mortality than with the other related death followed by other opportunistic infections and unknown infections but lower death from diarrhea as compared to other infections. After 65 months, unknown infections had higher risk death as compared to the other infections. This result indicated that the unknown infection should be differentiated, listed out and given due attention by health professionals. There is no significant difference in mortality risk from these causes of infections, since p-value (p= 0.025<0.05).

## Cause-specific hazard regression

Estimation of the risk of a male patient, age category 31-40 years and whose CD4 count is between 201-350 cells/mm^3^, the output shows that the cumulative incidences of death of this patient from other OIs at time points of 30, 50, 80 months were 0.31,0.37 and 0.40, respectively. The cumulative incidences of death of these patients from TB at the same time point were 0.28, 0.33 and 0.35. Unknown infection at time points of 30, 50, 80 months were 0.15, 0.19 and 0.20 respectively and diarrhea were 0.017, 0.019 and 0.020 respectively. Based on these numerical figures we can conclude that death due to opportunistic infection is more likely happened than the others causes.

## Sub-distribution hazards (SHs) model

Sub-distribution hazards (SHs) model is also known as Fine-Gray model. It is a Cox proportional regression model but the cumulative incidence is associated with SHs. The motivation for Fine-Gray is that the effect of a covariate on cause-specific hazard function may be quite different from that on Cumulative incidence function (CIF). In other words, a covariate may have strong influence on cause-specific hazard function, but have no effect on CIF[2]. The difference between cause-specific hazard and sub-distribution is that the competing risk events are treated differently. The former considers competing risk events as non-informative censoring, whereas the latter takes into account the informative censoring nature of the competing risk events[14].

### Prediction

Here we have considered four hypothetical individuals and using certain covariate combinations using the fitted Fine-Gray model selected based on the significance of the model. Age, sex, and starting CD4 levels are defined for individuals as covariates. The first individual, female, 29 years old and CD4 count is >=501. Referring with Fig 4, she is belonging to younger age but she is more likely to be died. This mean even if she had higher CD4 count, being a female was high risk of death early the start of ART.

**Fig 4.**
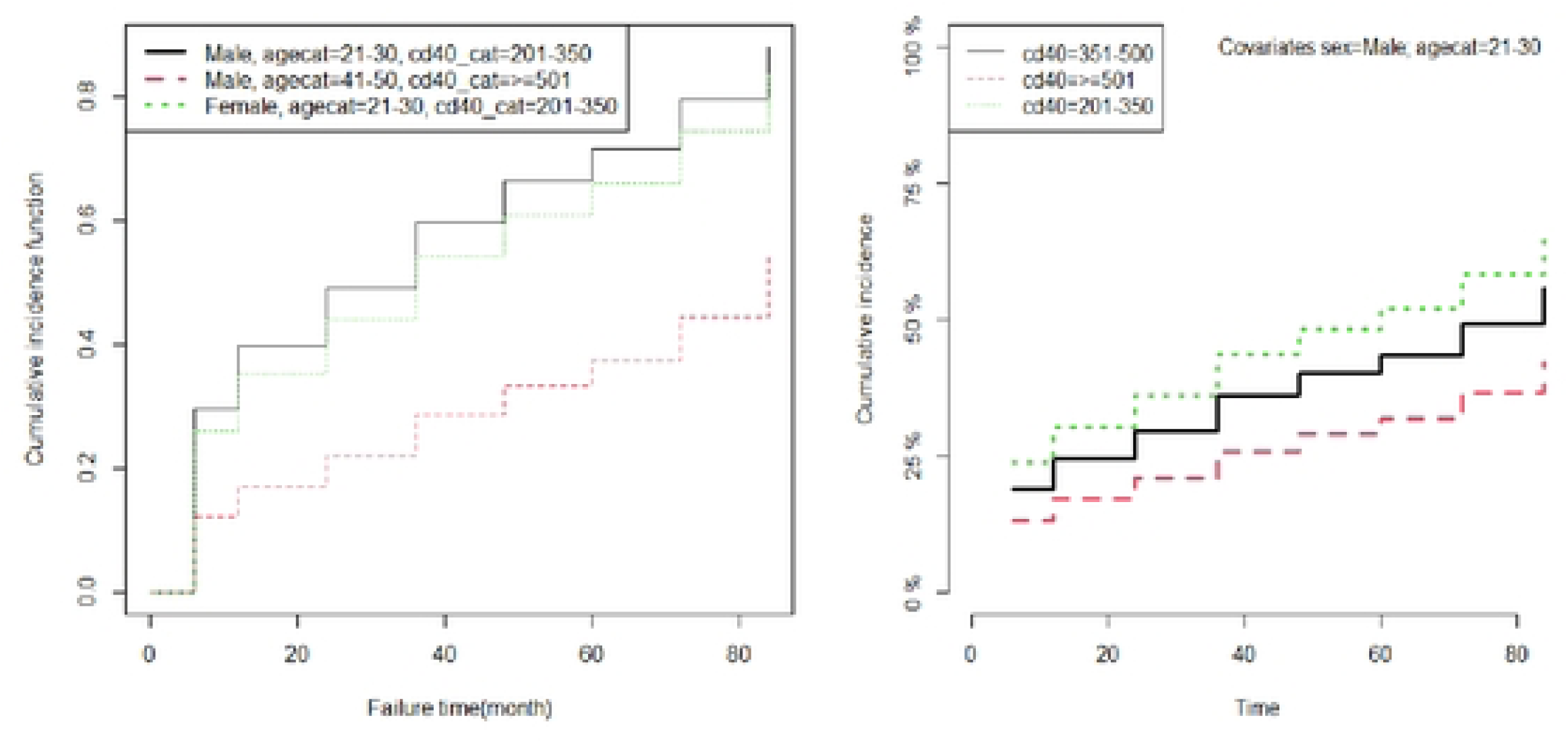
Title Parametric estimates of cumulative incidence functions for three patients with given covariate.

The second individual is a man who is 60 years old and his CD4 count is < 200. This 60 year old HIV patient has lower risk of death. This means, male patient has at lower risk than female during the first 3 years of the follow-up time.

Fig5 graphical output gives CIF for patients with characteristics specified in extracted data had three variables: Age, CD4 count, WHO clinical stage and Patient Residence. The link argument controls the link function to be used, “prop” for the Fine-Gray regression model with values age=52, CD4 count, WHO clinical stage and Residence compared in the four-graph panel with four of causes: tuberculosis, other opportunistic infection and diarrhea; and unknown causes. Then other opportunistic infection, tuberculosis, unknown and diarrhea causes were higher risk of death followed respectively. when we compared in each cause, those patients had higher WHO clinical stage higher risk as compare to other combination in the patient with the variable combination due to tuberculosis causes and other opportunistic but in contrast with tuberculosis cause the patient who had lower CD4 count high risk of death than the other combination. Blow Fig 5, as the baseline cd4 count increases, then decreases significantly the risk of death in all causes of death.

**Fig 5.**
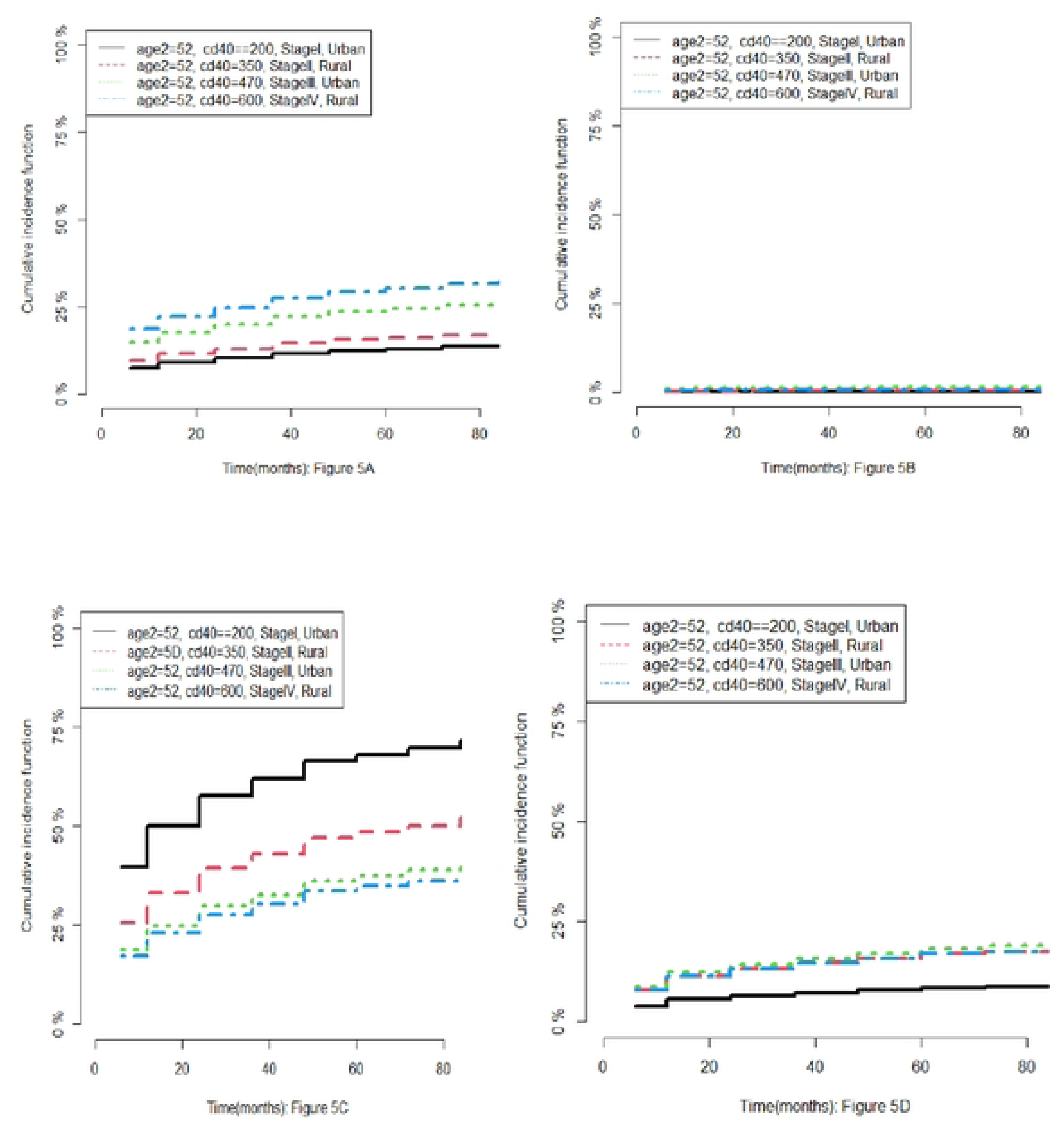
Title Estimated CIF. Fig 5 legend Figure 5A;TB, Figure 5B:Diarrhea, Figure5C:Other Infection and Figure 5D;Unknow Causes of Deaths of HIV/AIDS Patients by CD4 cell count using cause-specific hazard models.

#### Factors associated with opportunistic infections related mortality

In this study, we estimated the cumulative incidence of death from tuberculosis, diarrhea, other opportunistic diseases, and unknown infections using a cause-specific, sub-distribution, and flexible parametric hazard model. In univariate analysis, prognostic factors with p-values less than 0.2 were chosen, and the results are reported in Tables 2 & 3 below, and age group, patient residence, WHO stage, and baseline CD4 count met the criteria for multivariate analysis. Age group, WHO stage, residence and baseline CD4 count were statistically significant (P-value<0.05), Participant with age group 21-30 years had 1.78, 1.43 and 2.02 times higher risk of hazard of death due to tuberculosis causes as compared to age group <=20 years in the cause specific, sub-distribution and flexible parametric hazard model [HR= 1.78, 95% CI (1.24 - 2.541), SHR=1.43, 95% CI(.002 −1.98); HR=2.02, 95% CI(1.41-2.88)] respectively. But Death due to diarrhea cause the patient age group 40-50 years less like as compared to age group <20 years with HR =0.24 and 0.23 which slightly similar in the two models cause specific and flexible parametric hazard model [HR= 0.24, 95% CI (0.06-0.93), HR=0.23, 95% CI(0.06-0.92)] respectively but not significant in sub-distribution model. Since age is a confounding variable and majority of the patients 56% were died at early of the follow up period (6 months). The patient live in the rural area were higher risk death due to tuberculosis in the three models and nearly similar hazard ratio HR=1.37, 1.37 & 1.29 as compared to patients live in the Urban area in the cause specific, sub-distribution and flexible parametric hazard model [HR= 1.37, 95% CI (1.11-1.68), HR=1.37, 95% CI(1.13-1.65) and HR=1.29, 95% CI(1.04-1.58)] respectively. But patient live in rural area had 0.77, 0.76 and 0.78 less risk of death due to other opportunistic infection in almost similar in three models as compared to patients live in the urban area in the cause specific, sub-distribution and flexible parametric hazard respectively.

**Table 2:** Result of Cause specific Hazard Model and Sub-distribution Hazard Model.

**Table 3.** Result of Flexible Parametric Proportional Hazard Model.

The cause specific, sub-distribution and flexible parametric hazard model an increase in baseline CD4 count cells/mm3 is found to be strongly associated with the survival of HIV/AIDS patients and it is found to be a significant factor for the probability of death due to tuberculosis cause (P-value <0.05) .CD4 count category 351-500 count cells/mm3 less risk death due to tuberculosis than CD4 count category <200 count cells/mm3 with HR=0.58 and HR=0.54 in cause specific and flexible parametric hazard model (HR= 0.58, 95% CI (0.34, 0.98) and HR=0.54, 95% CI(0.32-0.92)] respectively. But sub-distribution was not significant death due to tuberculosis, diarrhea, other opportunistic infection and other unknown infection causes. WHO stage is found to be a significant factor for the probability of death due to any cause in the cause specific model (P-value <0.05).The result indicated that higher WHO stage nearly similar result in three models in the cause specific, sub-distribution and flexible parametric hazard ratio more than 2 times higher risk of death due to tuberculosis as compared to lower WHO stage. The reason WHO clinical stage increases the risk of death in all causes, the patient immunology decreases and they are exposed by opportunistic infectious

## Discussion

In this study, we used a national dataset that included all regional states and two administrative cities, and so it can give evidence to national health policy on the causes of mortality of HIV/AIDS patients under the ART category. It could be considered as an extension from the previous few studies already done in one region and health center from November 2019 to march 2020 patients under ART. We have discussed the three common methods for handling competing risks and their applications to regression setting.

The cause specific hazard model, sub-distribution hazard model and flexible parametric hazard model shows in the analysis output Table 2 & 3 produced nearly similar results with regard to the effect of covariates. Tuberculosis is the most common opportunistic infection cause of death among HIV/AIDS patients supported by previous studies [15–18]. During the period of observation, 34.7% were died due to tuberculosis, 44.7% were died due to other opportunistic infection but the others died due to diarrhea and unknown infection causes were 20.6%.This study agree with the study[15] and higher the study conducted in France[19]. According to all the methodologies, age group, CD4 count and WHO stage were significant indicators for HIV/AIDS patients.

Male patients had better survival than female HIV patients the start of ART treatment than female but female were better than male after 3 years of treatment periods [15]. Patient had nearly high risk of probability of death due to other opportunistic infection and Diarrhea in the first 2 years but probability of death due to unknown were lower as compared to the others causes. After 3 years, diarrhea had lower risk as compared to the others opportunistic infection and unknown infections and in agreement with the study[20] but in contrast with the study[21].

According to all the sub-distribution hazard models, flexible parametric and cause specific hazard models approaches, CD4 cells count is a key indicator for HIV/AIDS patients. Cause specific, sub-distribution and flexible parametric hazard ratios for tuberculosis deaths were found that the higher CD4 cell count cells/mm^3^ patients were less risk of death due to tuberculosis compared to the patients with lower CD4 cell count cells/mm^3^. This study was lower result than with the studies [15, 17].

The patient with higher age group were higher risk of death as compared to patient with lower age group and in agreement with the study [22]but death due to diarrhea were less as compared to lower age group this in agreement. This could be the cofounding effects of age.

The results of cause specific, sub-distribution and flexible parametric hazard model are found to be more than 2 times higher risk death in tuberculosis, diarrhea and opportunistic infection patient in ART treatment in WHO stage IV as compared to lower WHO stage I lower with the studies done previously[21]. Because of this, it may be wise on the side of treatment and care providers to focus on early detection of opportunistic infection and fast action should be taken to reduce morbidity and improve quality of life for persons with HIV/AIDS. Early detection and treatment of tuberculosis in HIV-infected people need to be prioritized. The prediction of patient risk with fewer combinations of variables weight, age and sex, CD4 cell count had lower risk as the patient had increase cd4 count.

There are several important benefits to this study. To our knowledge, this study is the first in Ethiopia compared death risks due to tuberculosis and other opportunistic infection in Ethiopian HIV/AIDS positive individuals. The overestimation of cumulative incidence and bias in the covariate effects might both be avoided. Second, our study had a sizeable sample of 39590 participants from across the country and a protracted follow-up period of (84 months). Last but not least, we looked into the connection between the time between starting ART and mortality in HIV/AIDS patients with high and low CD4+ cell counts.

According to the current study findings, late HIV/AIDS diagnosis at an older age, in stages 3 or 4, or with a lower CD4 count may raise the risk of death considerably. These findings were consistent with those of other investigations[15, 21].

This study has some limitations, the dynamic fluctuations in CD4+ cell counts over time and causes of death record not appropriate this could affect to see the survival of HIV/AIDS patients. Due to the study’s record review methodology, some variables including height and hemoglobin were missing. Due to the incompleteness of the data, BMI and anemia status were excluded from the current analysis.

## Conclusion

The results of this study clearly shown that tuberculosis, diarrhea, opportunistic infection and unknown cause are substantially accelerating death for HIV/AIDS patients in Ethiopia. Applying effective strategies is needed to achieve on time diagnosis of individuals with HIV and provide them with HIV/AIDS care and treatment services to enhance the survival of the patients. Moreover, female younger age living with HIV/AIDS requires more attention when receiving HIV/AIDS care and treatment as they experience a higher risk of death the first 3 years after ART initiation. The findings of the current research revealed that early HIV/AIDS care and treatment could substantially reduce opportunistic infection death among HIV/AIDS patients. Therefore, to reduce mortality rate HIV/AIDS patient, the current strategies should be revised to improve the timing of treatment initiation and also to optimize the adherence to the treatment.

## Abbreviations

ART: Antiretroviral therapy
HAART: Highly active antiretroviral therapy
AHRI: Armauer Hansen Research Institute
AIDS: Acquired Immunodeficiency Syndrome
CD4: Cluster of Differentiation 4 or Classification Determinant
FHAPCO: Federal HIV/AIDS Prevention & Control Office
HIV: Human Immunodeficiency Virus
NAPES: National ART Program Effectiveness Study
IRB: Institutional Review Board

## Ethics approval and consent to participate

Ethical clearance was initially obtained back in June/July 2019 from ALERT/Armauer Hansen’s Research Institute’s (AHRI’s) IRBs [P048/18]. The need for consent was waived by the above mentioned ethics review committee as the study is based on a secondary data(record review). Consecutively, before dispatching the team to the different regions, a formal official approval and support letter describing the purposes of the study was written from Federal MoH to the respective Regional health bureaus and in turn to each zonal offices and hospitals and then from Zonal Health Bureaus to the Woreda Health Offices before reaching the health facilities.

## Consent for participation (informed consent)

Since the study design was retrospective and the data obtained from record review and informed consent is not applicable (N/A).

## Availability of data and materials

The data sets analyzed in this study available from the corresponding author on reasonable request. The R code used to analyze the data provided as a supplement of the article.

## Authors’ contributions

Tsegaye H Kumsa and Zeytu G Asfaw generated the idea, the corresponding author THK did the data analysis and interpretation and ZGA played an advisory role. THK and Andargachew M ulu (AM) have generated the data and Andargachew Mulu were the project investigator. Adane Mihret reviewed the manuscript. All authors read and approved the final manuscript.

## Acknowledgements

We would like to also express our gratitude to MoH, Regional, Zonal and Distract Health Authorities for the administrative effort made for the data collectors. We are also thankful for the collaboration effort both from Armauer Hansen Research Institute, Federal HAPCO and Ministry of Health and Regional and City Administration Health Bureaus. We thank also the data management team, finance and procurement, general service and the entire AHRI top and middle level management team for the integrated service and leadership that they provided for the study team during the data collection

**Authors’ information (optional)**

